# Re-emergence of Sylvatic Dengue 2 during an outbreak in Southeastern Senegal, Kedougou 2020

**DOI:** 10.1101/2023.11.22.23298824

**Authors:** Idrissa Dieng, Maryam Diarra, Bacary Djilocalisse Sadio, Alioune Gaye, Bocar Sow, Marie Henriette Dior Ndione, Diawo Diallo, Mignane Ndiaye, Amadou Diallo, Safietou Sankhe, Martin Faye, Boly Diop, Amadou Alpha Sall, Gamou Fall, Oumar Faye, Cheikh Loucoubar, Ousmane Faye, Scott C. Weaver, Mawlouth Diallo, Mamadou Aliou Barry, Moussa Moise Diagne

## Abstract

Dengue outbreaks in West Africa, linked to urban cycle viruses, pose a significant public health threat. In 2020, a sylvatic Dengue 2 outbreak in Kedougou, southeastern Senegal, resulted in 59 confirmed cases, suggesting these strains may not require additional adaptation but could re-emerge into urban transmission cycles in the region.

## The study

Kedougou, Senegal’s southeastern region, is a significant arbovirus hotspot (*1,2*) with decades of comprehensive surveillance through both a nationwide Syndromic Sentinel Surveillance network (*3*) and a passive surveillance in several public health structures in Kedougou and Saraya districts (*2*). Whole blood samples collected in the different healthcare sites are sent to the WHO collaborating Center for Arboviruses and Hemorrhagic Fever Viruses in IPD in Dakar for laboratory analysis, as previously described (*2,4*).

A old man in their 20s with an arboviral infection syndrome was admitted to a health care center in November 2020. The RNA was amplified using a pan-Dengue (panDENV) one-step RT-qPCR assay (*4*), confirming a dengue infection. The arbovirus surveillance reported 36 additional cases, including 27 RT-qPCR positive samples. An investigation team from both the Ministry of Health and IPD was mobilized in December 2020, identifying 14 recently infected people out of 42 suspected cases through the retrospective tracing of visit records in health centers. From December 2020 to late January 2021, four more RT-qPCR positive cases and four serological confirmed ones were reported through passive surveillance.

A working case definition was developed as previously described (*5*) for i) suspected cases, referring to sudden onset of fever with arbovirus symptoms, and ii) confirmed cases, indicating infection confirmed by laboratory methods. Door-to-door case research was conducted in housing areas, collecting socio-demographic and clinical data. Continuous variables were summarized as means or as median while dichotomous or categorical variables were described by percentages with 95% confidence interval as previously described (*3*); the Kruskal–Wallis test was used to compare the mean age between negative and confirmed dengue cases. Pearson’s χ2-test or Fisher’s exact test were used to compare proportions between categories. A p-value < 0.05 was considered statistically significant. The statistical analysis was performed with Stata version15 software (StataCorp, LLC, College Station, TX, USA).

Until February 27, 2021, a total of 300 serum samples was collected from different localities across Kedougou region (figure 1). Overall, DENV infection was 19.6% (59/300) with 95%CI (15.1 – 24.2) corresponding to 32 RT-qPCR and 27 IgM positive cases (table 1). Among confirmed cases, the highest number was recorded in Saraya health district (n = 18) followed by Bandafassi PHC (n = 14), Kedougou health district (n = 14) and Military Camp (n = 13) (figure 1).

**Table 1:**
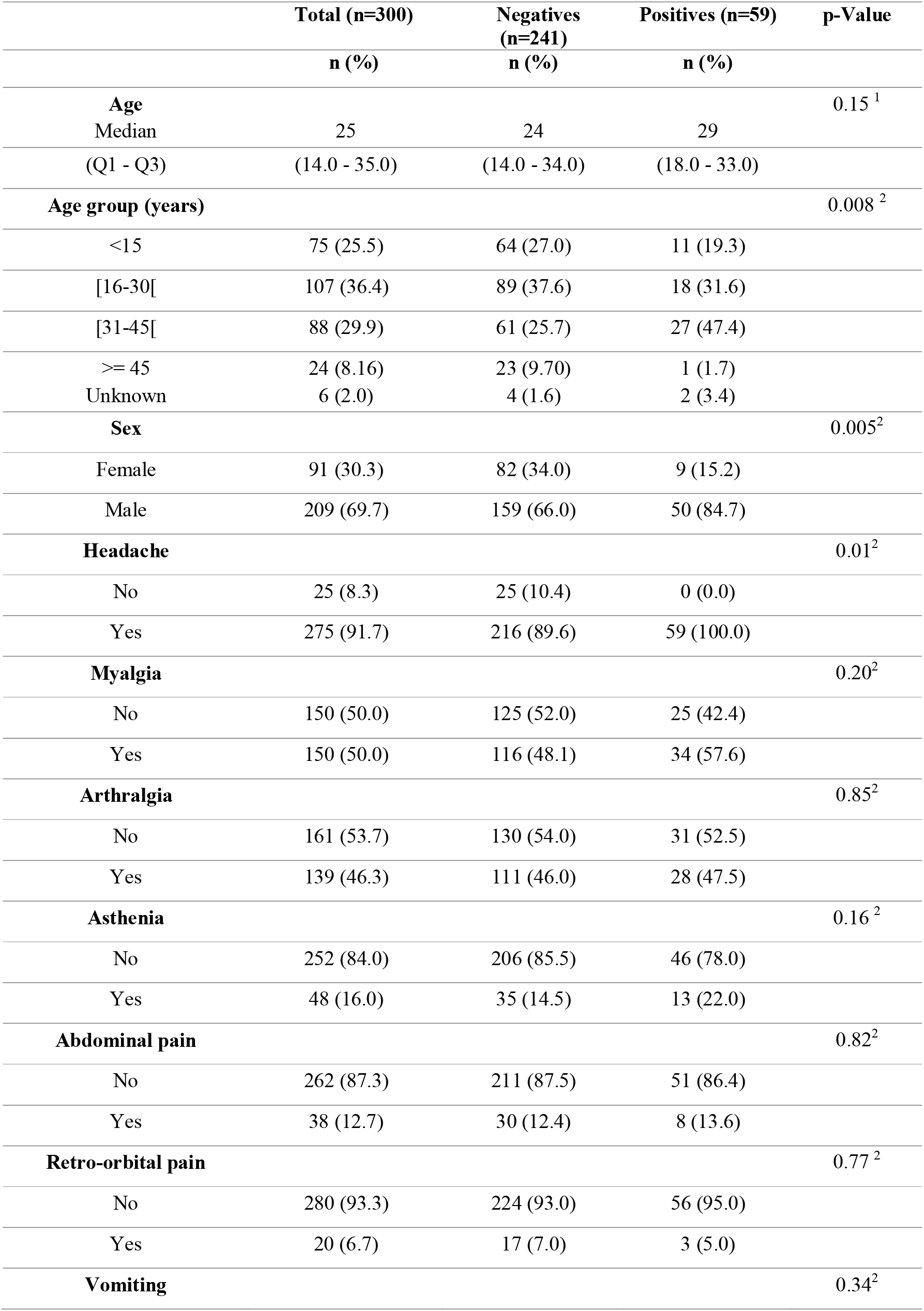
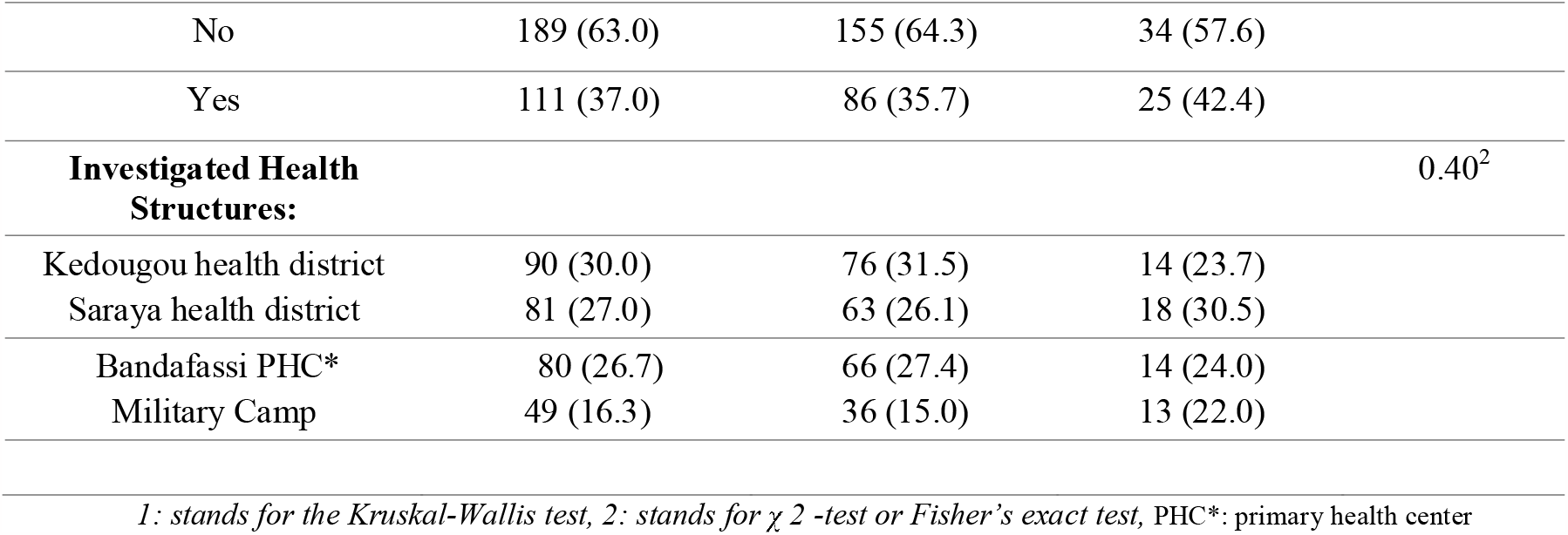
Epidemiological and clinical characteristics of suspected and confirmed dengue fever cases investigation, Kedougou, Senegal, 2020

**Figure 1:**
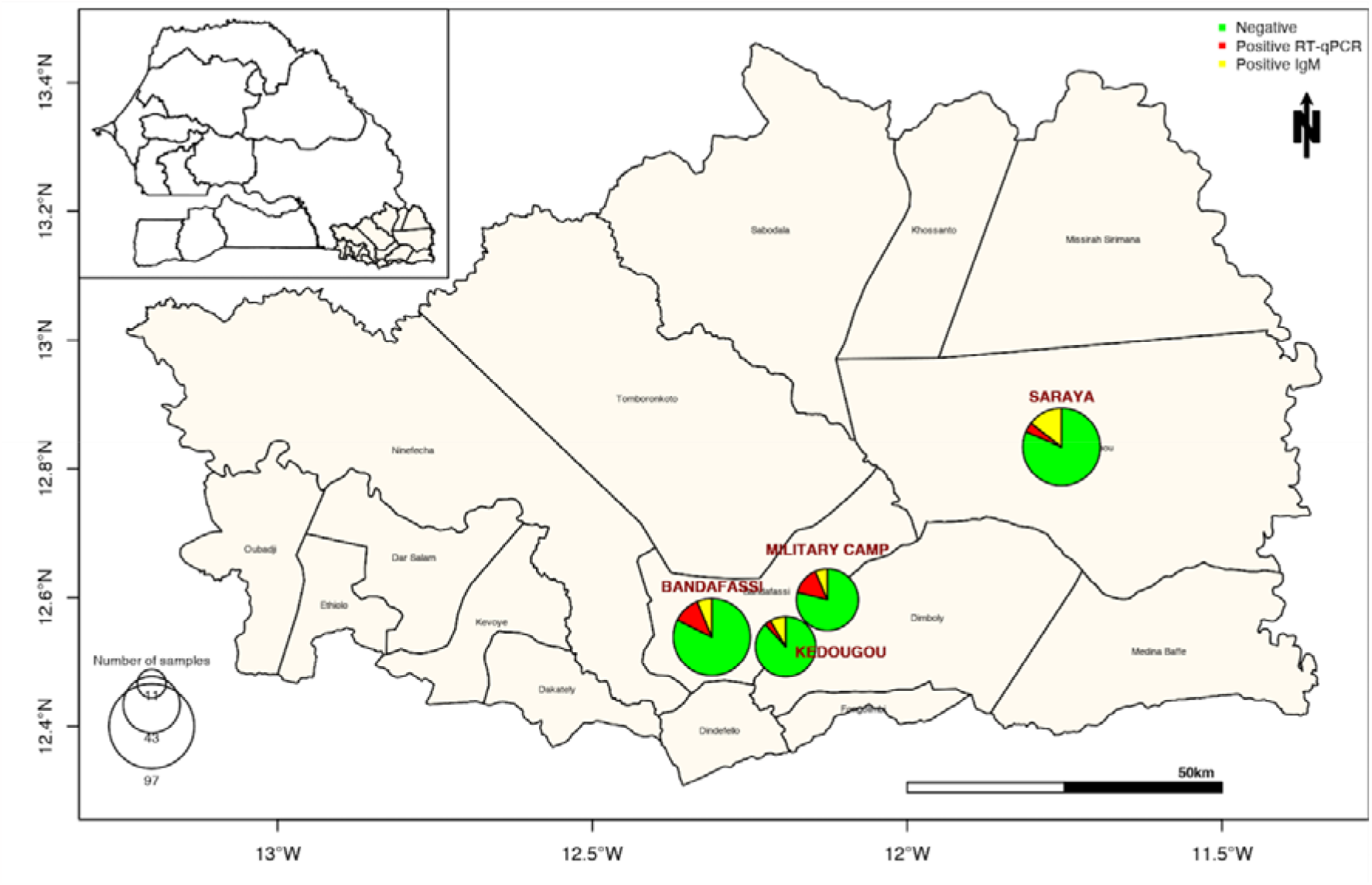
Spatial distribution of reported cases in the sylvatic Dengue outbreak in Kedougou region between November 2020 to February 2021.

Men were more affected than women, with a sex ratio of 5.5 for confirmed cases (p = 0.005; Pearson’s χ2-test). The mean age of all patients was 25.5 years (SD: ±13.8 years), with most cases falling in the 30-45 age group (47.4%), followed by the 15-30 age group (31.6%), and the 45+ age group (1.7%); the positivity rate varied significantly according to the age group (p=0.008; Pearson’s χ2-test). The most common symptoms reported were headaches significantly recorded in all confirmed dengue cases (p=0.01), followed by myalgia (57.6%) and arthralgia (47.5%) (table 1).

Beside human investigation, entomological surveillance from August to November 2020 in 50 sites across five land cover classes (forest, barren, savannah, agricultural lands and villages) collected 15,937 mosquitoes, including over 56 species in 7 genera. Over half were known sylvatic or peridomestic DENV vectors (*6*) (table 2). Interestingly, no DENV was identified in monospecific mosquito pools, while a concomitant Yellow fever virus circulation was detected as previously reported (2).

**Table 2:** Mosquito species collected from August to November, 2020 in Kedougou region.

| Species | Number of individuals (%) |
| --- | --- |
| *Aedes dalzieli* | 3559 (22.3 %) |
| *Aedes furcifer* | 2332 (14.6 %) |
| *Aedes aegypti* | 1298 (8.4 %) |
| *Aedes vittatus* | 971 (6.1 %) |
| *Aedes luteocephalus* | 766 (4.8 %) |
| *Aedes taylori* | 330 (2.1 %) |
| *Aedes africanus* | 279 (1.8 %) |
| Others | 6402 (40.2%) |
| Total | 15937 (100%) |

A molecular serotyping assay (*7*) was undertaken on the panDENV RT-qPCR positive human samples with no success, suggesting that the strain may belong to the DENV-2 sylvatic genotype, according to a previous work (*8*).

Samples were then sequenced by an amplicon-based approach on a MinION MK1C sequencer (Oxford Nanopore Technologies, UK) using two separate pools of a sylvatic DENV-2 specific primer scheme amplifying the entire coding region of the genome.

Libraries were prepared using the Rapid Barcoding Kit RBK 110.96 (ONT, UK) and load onto a R9 flow cell. Data analysis was performed as previously described (*8*) and consensus genomes were aligned with a representative dataset of available DENV-2 sequences using MAFFT (*9*). A Maximum likelihood tree was built using IQ-TREE (*10*). Phylogenetic analysis confirmed that sequenced strains belong to the sylvatic DENV-2 genotype and are closely related to a strain identified from a traveler returning from Guinea-Bissau in 2009 *(11)* (Figure 2). In 2021, a sylvatic DENV-2 infection was reported in Senegal in Kolda, in the south Senegal, near the border with Guinea-Bissau (*8*).

**Figure 2:**
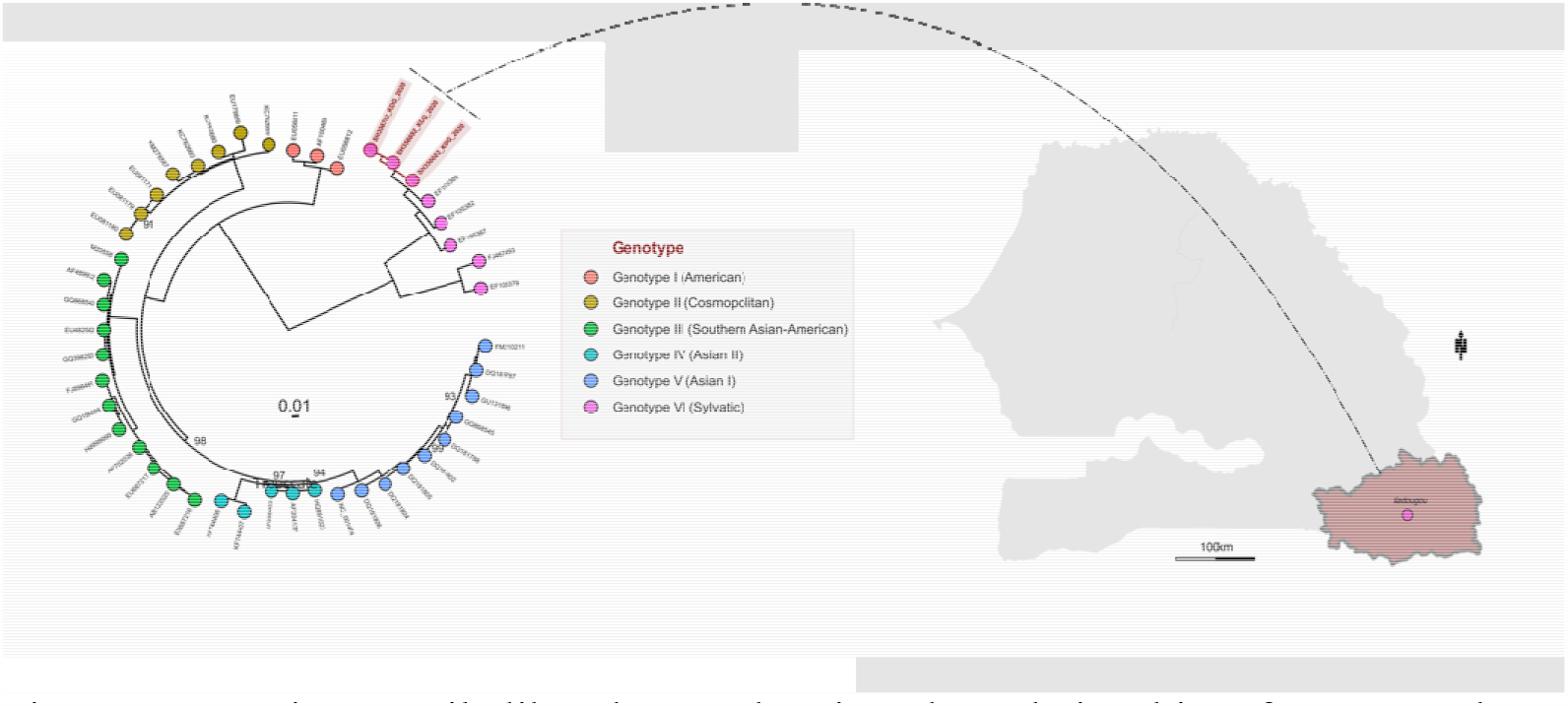
Maximum Likelihood tree showing the relationship of sequenced DENV-2 during outbreak in Kedougou shows that sequenced strains belong to the sylvatic genotype of DENV-2 and are closely related to sequence obtained in 2009 in Guinea-Bissau.

The study reveals that while the epidemic DENV in Senegal is multi-serotypes (*12*), sylvatic strains are still circulating and can cause large outbreaks. This supports previous research suggesting that sylvatic strains in humans may not require additional virus adaptation (*13*) but could re-emerge into urban transmission cycles. These strains should be considered as agents with epidemic potential, especially in areas like Kedougou where the ecosystem combines human and non-human primates and primatophilic mosquitoes (*6, 14*). Large-scale genomic surveillance is needed and molecular diagnostic tools should be updated.

## Data Availability

All data produced in the present study are available upon reasonable request to the authors

## Acknowledgments

We would like to convey special thanks to the virology lab workers at the Institut Pasteur de Dakar.

## References

1. Sow, A.; Loucoubar, C.; Diallo, D.; Faye, O.; Ndiaye, Y.; Senghor, C.S.; Dia, A.T.; Faye, O.; Weaver, S.C.; Diallo, M.; et al. Concurrent malaria and arbovirus infections in Kedougou, southeastern Senegal. Malar. J. 2016, 15, 47.

2. Diagne, M.M.; Ndione, M.H.D.; Gaye, A.; Barry, M.A.; Diallo, D.; Diallo, A.; Mwakibete, L.L.; Diop, M.; Ndiaye, E.H.; Ahyong, V.; et al. Yellow Fever Outbreak in Eastern Senegal, 2020–2021. Viruses 2021, 13, 1475.

3. Barry, M.A.; Arinal, F.; Talla, C.; Hedible, B.G.; Sarr, F.D.; Ba, I.O.; Diop, B.; Dia, N.; Vray, M. Performance of case definitions and clinical predictors for influenza surveillance among patients followed in a rural cohort in Senegal. BMC Infect. Dis. 2021, 21, 31.

4. Sow, A.; Faye, O.; Diallo, M.; Diallo, D.; Cheng, R.; Faye, O.; Diagne, C.T.; Guerbois, M.; Weidmann, M.; Ndiaye, Y.; et al. Chikungunya Outbreak in Kedougou, Southeastern Senegal in 2009–2010. Open Forum Infect Dis. 2018, 5.

5. Dieng, I.; Barry, M.A.; Talla, C.; Sow, B.; Faye, O.; Diagne, M.M.; Sene, O.; Ndiaye, O.; Diop, B.; Diagne, C.T.; Fall, G.; Sall, A.A.; Loucoubar, C.; Faye, O. Analysis of a Dengue Virus Outbreak in Rosso, Senegal 2021. Trop. Med. Infect. Dis. 2022, 7, 420. 10.3390/tropicalmed7120420

6. Diallo, M., Ba, Y., Sall, A. A., Diop, O. M., Ndione, J. A., Mondo, M., Girault, L., Mathiot, C. Amplification of the sylvatic cycle of dengue virus type 2, Senegal, 1999-2000: entomologic findings and epidemiologic considerations. Emerg Infect Dis, 2003, 9, 3.

7. Dieng, I.; Cunha, M.; Diagne, M.M.; Sembène, P.M.; Zanotto, P.M.A., Faye, O.; Faye, O.; Sall, A.A. Origin and Spread of the Dengue Virus Type 1, Genotype V in Senegal, 2015-2019. Viruses, 2021, 13, 1.

8. Dieng, I.; Sagne, S.N.; Ndiaye, M.; Barry, M.A.; Talla, C.; Mhamadi, M.; Balde, D.; Toure, C.T.; Diop, B.; Sall, A.A.; et al. Detection of human case of dengue virus 2 belonging to sylvatic genotype during routine surveillance of fever in Senegal, Kolda 2021. Front. Virol, 2022

9. Katoh, K.; Rozewicki, J.; Yamada, K.D. MAFFT online service: multiple sequence alignment, interactive sequence choice and visualization. Brief Bioinform. 2019, 20, 4.

10. Nguyen, L.T.; Schmidt, H.A.; von Haeseler, A.; Minh, B.Q. IQ-TREE: a fast and effective stochastic algorithm for estimating maximum-likelihood phylogenies. Mol Biol Evol. 2015, 32, 1.

11. Franco, L.; Palacios, G.; Martinez, J. A.; Vázquez, A.; Savji, N.; De Ory, F.; Sanchez-Seco, M.P.; Martín, D.; Lipkin, W.I.; Tenorio, A. First report of sylvatic DENV-2-associated dengue hemorrhagic fever in West Africa. PLoS Negl Trop Dis, 2011, 5, 8.

12. Dieng, I.; Ndione, M.H.D.; Fall, C.; Diagne, M.M.; Diop, M.; Gaye, A.; Barry, M.A.; Diop, B.; Ndiaye, M.; Bousso, A.; et al. Multifoci and multiserotypes circulation of dengue virus in Senegal between 2017 and 2018. BMC Infect Dis, 2021, 21, 1.

13. Vasilakis, N.; Holmes, E,C.; Fokam, E,B.; Faye, O.; Diallo, M.; Sall, A.A.; Weaver, S.C. Evolutionary processes among sylvatic dengue type 2 viruses. J Virol, 2007, 81, 17.

14. Diallo, D., Chen, R., Diagne, C. T., Ba, Y., Dia, I., Sall, A. A., Weaver, S. C., Diallo, M. Bloodfeeding patterns of sylvatic arbovirus vectors in southeastern Senegal. Trans R Soc Trop Med Hyg, 2013, 107, 3.

